# Genotype Variants and Other Covariates are not significantly associated with Malaria Positivity Status among Pregnant Women visiting Two Public Health Facilities in Southwest, Nigeria

**DOI:** 10.1101/2025.09.18.25336062

**Authors:** O.R. Oyerinde, G.A. Adegbite, O.O. Ayodele, F.J. Femi-Olabisi, O.G. Ademowo

## Abstract

Malaria remains a disease of endemicity in Sub-Saharan African countries but most particular in Nigeria. Pregnant women are among the vulnerable group of individuals most affected by the disease. However, previous studies focused on the effect of genotype variants and other covariates on malaria status with emphasis on the entire population regardless of vulnerability levels. Hence as part of the effort in controlling malaria within a particular malaria-vulnerable group (e.g. pregnant women), this study investigates the effect of genotype variants, weight, height and temperature on malaria status.

A total of seven hundred and seven (707) pregnant women were recruited from two government healthcare facilities in Ibadan in Nigeria, for this study after necessary ethical approval and consent were obtained. Binary Logistic Regression was carried out using malaria positivity rate as dependent variable while genotype variants (AA, AC, AS, SC, SS), weight, height and temperature were the independent variables. The odd ratios were obtained for these independent variables as well as their level of statistical significance (p-values) in relation to malaria status.

The odds ratio for genotypes AA, AC, AS, SC and SS of being malaria positive were 1, 0.3, 0.04, -14 and -14, respectively. The odds ratio for weight, height and temperature to influence malaria positivity status were -0.01, -0.02 and 0.04, respectively. However, genotype variants, weight, height and temperature were not significantly associated with malaria positivity status.

These findings demonstrate that the only effective way of preventing malaria for pregnant women regardless of their genotype and other covariates, is to ensure the correct use of long lasting insecticidal treated nets and indoor residual spray. Also, all pregnant women should ensure that they carry out required antenatal care visits, that will include the administration of Intermittent Preventive Treatment in Pregnancy.

## Introduction

Malaria remains a disease of endemic burden in Nigeria, one of the Sub-Saharan African countries. (Mezieobi *et al*., 2025, Ogbozor, 2025; Eneh *et al*., 2025). Over the years, several studies, both experimental and *in silico* studies have shown that pregnant women and under-five children are the most affected by the disease and can be affected by severe cases of the disease (Adegbite *et al*., 2022, Zegeye *et al*., 2025). Hence, national malaria control programs in Nigeria and some other malaria-affected countries have followed several stipulated guidelines from the World Health Organization in safeguarding the lives of the children and pregnant women (de Cola et al., 2022, Kazanga *et al*., 2025).

Seasonal Malaria Chemoprevention and Intermittent Preventive Treatment in Pregnancy (IpTp) are the commonest tailored interventions for children and pregnant women, respectively (de Cola *et al*., 2022, Kazanga *et al*., 2025). IpTp is often administered during antenatal care visits (Kretchy *et al*., 2025). Specifically, the use of either long lasting Insecticidal Treated Nets or Indoor Residual Spray has been recommended for the general population regardless of the degree of malaria vulnerability (Adegbite *et al*., 2023). Although malaria vaccines are also gradually gaining momentum towards full acceptance and adoption (Alum *et al*., 2025).

However, several schools of thought have proposed that different individual characteristics can affect disease vulnerability (Adegbite *et al*., 2022). Genotype variants (AA, AS, AC, SC, SS), weight, height, temperature and others, can affect malaria positivity status (Raballah *et al*., 2025). However, the focus of most of these studies are on the general population, with less emphasis on the identified most malaria-vulnerable individuals (i.e. children and pregnant women). Hence, this work investigated the effect of genotype, weight, height and temperature on malaria positivity rate using a machine learning method on one of the vulnerable groups (i.e. pregnant women).

Machine learning methods are advanced statistical models, relevant in the field of data science, in discovering new patterns from data (Mthethwa, & Melesse, 2025). Machine learning models’ earliest examples include linear, non-linear, binary, multinomial logistic regression methods (Mthethwa, & Melesse, 2025). Linear regression could be classified as simple or multiple (Mthethwa, & Melesse, 2025). Simple linear regression involves a representative linear relationship, between one dependent numerical variable and one independent variable (Mthethwa, & Melesse, 2025). Multiple linear regression involves a representative linear relationship, between one dependent numerical variable and two or more independent variables (Mthethwa, & Melesse, 2025). Nonlinear regression centred around the concept of curve fitting i.e. fitting data to models using mathematical functions e.g. logarithmic, exponential, sine among others (Mthethwa, & Melesse, 2025). Maximum Likelihood Estimator and Non-Least Square methods are common functions implemented in computer program libraries in carrying out curve fitting (Mthethwa, & Melesse, 2025).

Binary logistic regression involves a dependent categorical variable that can have only two possible responses i.e. yes or no; in relation to other independent variables that can be numerical and/or categorical (Mthethwa, & Melesse, 2025). Multinomial logistic regression involves a dependent categorical variable that can have more than two possible responses; in relation to other independent variables that can be numerical and/or categorical (Mthethwa, & Melesse, 2025). Due to the peculiarity of this work and its method fit, this study used a binary logistic regression model to statistically analyse the relationship between malaria positivity/negativity status in relation to the genotype variants, weight, height and temperature among pregnant women visiting two public healthcare facilities in Ibadan, Southwestern part of Nigeria.

## Materials and Methods

### Ethical Considerations

Ethical approval for this work was gotten from the University College Hospital Ethical Review Committee of the University of Ibadan in Nigeria. Further approval was obtained specifically from the Oyo State Ministry of Health. Informed consent was obtained from each pregnant woman enrolled for this study, prior to participation.

### Study Site

This study was carried out in two government-based Healthcare Facilities (including Agbongbon Primary Healthcare Centre and Adeoyo Maternity Hospital) in Ibadan, Southwest Nigeria. These healthcare facilities serve the local inhabitants who are mostly ‘home makers and traders. The average attendances at each ante-natal clinics visit of these hospitals are about 120 and 25 pregnant women respectively.

### Data Collection

707 pregnant women were recruited for this study from the two healthcare facilities. Through a Likert Scale approach, the recruited pregnant women were interviewed by trained data collectors. For ease of communication, participants were interviewed via the local language of the Southwestern Nigeria which is Yoruba. Based on these interactions, basic sociodemographic characteristics of the respondents were gotten. Also, medical personnels from the hospitals help in taking vital signs and other relevant individual metrics, from the recruited pregnant women such as temperature, weight, height among others. Also, genotype tests were carried out for the pregnant women.

### Data Preprocessing and Analysis

The data collected contained some missing values. Imputation was carried out to replace missing values for weight, height, temperature and genotype variants. The malaria status (positive/negative) was the dependent variable while genotype variants (AA, AS, AC, SS, SC), weight, height and temperature were the independent variables. The analysis for our work was done using binary logistic regression implemented using the R programming language on the RStudio Integrated Development Environment. Odd ratios and associated p-values to demonstrate statistical significance were gotten for the independent variables in relation to malaria positivity status.

## Results and Discussion

Table 1 shows the odds ratio and associated p-values for the independent variables in influencing malaria positivity status. The odds ratio for genotypes AA, AC, AS, SC and SS of being malaria positive were 1, 0.3, 0.04, -14 and -14, respectively. The odds ratio for weight, height and temperature to influence malaria positivity status were -0.01, -0.02 and 0.04, respectively. However, genotype variants, weight, height and temperature were not significantly associated with malaria status.

| Malaria Positivity Status | Odds Ratio | P-Values |
| --- | --- | --- |
| Genotype AA | 1 | >0.05 |
| Genotype AC | 0.3 | >0.05 |
| Genotype AS | 0.04 | >0.05 |
| Genotype SC | -14 | >0.05 |
| Genotype SS | -14 | >0.05 |
| weight | -0.01 | >0.05 |
| height | -0.02 | >0.05 |
| temperature | 0.04 | >0.05 |

## Conclusion

This work used a binary logistic regression model to assess the statistical significance of genotype variants and other covariates on malaria positivity status. Based on findings from this study, there is no statistically significant importance of genotype variants and other covariates on malaria positivity status. Hence, this work opined that the only effective way of preventing malaria for pregnant women regardless of their genotype and other covariates, is to ensure the correct use of long lasting insecticidal treated nets and indoor residual spray. Also, all pregnant women should ensure that they carry out required antenatal care visits, that will include the administration of Intermittent Preventive Treatment in Pregnancy.

## Data Availability

Data used in this work is available upon reasonable request to the authors.

## Acknowledgements

The field, laboratory staff and recruited/consented participants are duly acknowledged for their role in the study.

## References

Adegbite, G., Edeki, S., Isewon, I., Dokunmu, T., Rotimi, S., Oyelade, J., & Adebiyi, E. (2022,). Investigating the epidemiological factors responsible for malaria transmission dynamics. In IOP Conference Series: Earth and Environmental Science (Vol. 993, No. 1, p. 012008). IOP Publishing.

Adegbite, G., Edeki, S., Isewon, I., Emmanuel, J., Dokunmu, T., Rotimi, S., … & Adebiyi, E. (2023). Mathematical modeling of malaria transmission dynamics in humans with mobility and control states. Infectious Disease Modelling, 8(4), 1015–1031.

Alum, E. U., Ainebyoona, C., Egwu, C. O., Onohuean, H., Ugwu, O. P. C., Uti, D. E., … & Echegu, D. A. (2025). Mitigation of Malaria in Sub-Saharan Africa through Vaccination: A Budding Road Map for Global Malaria Eradication. Ethiopian Journal of Health Sciences, 35(3).

de Cola, M. A., Sawadogo, B., Richardson, S., Ibinaiye, T., Traoré, A., Compaoré, C. S., … & Okell, L. C. (2022). Impact of seasonal malaria chemoprevention on prevalence of malaria infection in malaria indicator surveys in Burkina Faso and Nigeria. BMJ global health, 7(5), e008021.

Eneh, S. C., Obi, C. G., Ekwebene, O. C., Edeh, G. C., Awoso, O., Udoewah, S. A., et al. (2025). Eliminating malaria in Nigeria: insights from Egypt’s success and pathways to sustainable eradication. Malaria Journal, 24(1), 183.

Kazanga, B., Ba, E. H., Legendre, E., Cissoko, M., Fleury, L., Bérard, L., … & Landier, J. (2025). Impact of seasonal malaria chemoprevention timing on clinical malaria incidence dynamics in the Kedougou region, Senegal. PLOS Global Public Health, 5(1), e0003197.

Kretchy, I. A., Atobrah, D., Adumbire, D. A., Ankamah, S., Adanu, T., Badasu, D. M., & Kwansa, B. K. (2025). Enhancing the uptake of intermittent preventive treatment for malaria in pregnancy: a scoping review of interventions and gender-informed approaches. Malaria Journal, 24(1), 49.

Mezieobi, K. C., Alum, E. U., Ugwu, O. P. C., Uti, D. E., Alum, B. N., Egba, S. I., & Michael, E. C. (2025). Economic burden of malaria on developing countries: A mini review. Parasite Epidemiology and Control, e00435.

Mthethwa, S. M., & Melesse, S. F. (2025). A Machine Learning Approach to the Prediction of Malaria in Under-five Children: Analysis of the 2021 Nigerian Malaria Indicator Survey. The Open Public Health Journal, 18(1).

Ogbozor, I. B. (2025). Addressing sub-Saharan Africa’s burden of endemic diseases: a case for mandatory citizen participation in research on malaria and type 2 diabetes (Doctoral dissertation, Memorial University of Newfoundland).

Raballah, E., Anyona, S. B., Osata, S. W., Wasena, S. A., Onyango, C., Hurwitz, I., … & Perkins, D. J. (2025). Impact of age, HIV1, sickle-cell genotypes, and interferon-gamma gene upstream variants on malaria disease outcomes in a longitudinal pediatric cohort. Scientific Reports, 15(1), 13043.

Zegeye, A. F., Wassie, M., Tamir, T. T., Tekeba, B., Mekonen, E. G., Zeleke, G. A., & Gebrehana, D. A. (2025). Malaria-anemia comorbidity and its determinants among pregnant women in high-and moderate-malaria-risk countries in Sub-Saharan Africa. Infectious Diseases of Poverty, 14(1), 86.

